# Finding Functionality: Rasch Analysis of the Functionality Appreciation Scale in community-dwelling adults including adults with spinal cord injury and neuropathic pain in the US

**DOI:** 10.1101/2022.07.16.22277712

**Authors:** Sarah Feng, Sydney McDaniel, Ann Van de Winckel

## Abstract

The Functionality Appreciation Scale (FAS) measures an individual’s appreciation for the functions their body can perform, regardless of the individual’s physical limitations. To facilitate the use of this tool by researchers and clinicians, it is necessary to understand what ‘normal’ scoring looks like in healthy adults, as well as validate the scoring of this scale. We analyzed the FAS using Rasch Measurement Theory. FAS responses, demographic data, and clinical questionnaires were collected by the Brain Body Mind Lab (University of Minnesota) from 567 community-dwelling adults recruited at the Minnesota State Fair, including 14 adults with spinal cord injury. We evaluated item and person fit, targeting, unidimensionality, person separation reliability (PSR), local item dependence (LID), and differential item functioning (DIF) for demographic and clinical characteristics. We found a significant ceiling effect (26.98%) and a positive person mean location of 3.28±2.10 logits, indicating the FAS is too easy for the community-dwelling adults in the US. PSR was 0.79, indicating a capacity to differentiate groups of people according to their ability level of functional appreciation. We also compared the person mean location of adults who did or did not participate in body awareness training. Adults who practiced body awareness training had a higher person mean location (4.28, Interquartile Range [IQR] = 3.96 logits) than those who did not (2.73, IQR = 3.34), indicating a higher functionality appreciation. Overall, FAS demonstrated good item and person fit, but the current version of FAS will require more difficult items to be added to improve the targeting of the scale for use in a general population.

## Introduction

Functionality appreciation is characterized by the acknowledgment and value of everything the body is capable of doing, from communication to physical capacities (Alleva & Tylka, 2021). As research on the effects of mindfulness and other mind and body approaches are increasingly being used for treating chronic pain, mental health treatment, rehabilitation from injury, and other health conditions, it has become clear that body awareness and body image play a role in overall mental wellness (de Jong et al., 2016). The appreciation of body functionality has also shown a positive correlation with improved body image, establishing itself as a major facet of the construct (Alleva et al., 2017).

Previous studies on body functionality have sought to define the concept under limited terms of which bodily systems may qualify as “functional.” However, there are various forms of body functionality, specifically relating to internal functions and external functions. Some internal examples include digestion, the senses of sight or smell, and creativity. External functions include interaction with others, physical capabilities in movement, and hygienic practices (Alleva & Tylka, 2021). In a more inclusive sense, body functionality acknowledges the way the body functions according to its ability to accomplish specific needs, as opposed to limiting the definition of functionality to carrying out these processes in a particular way (Alleva & Tylka, 2021).

Most researchers have assessed functionality appreciation with surveys rather than physical activity, as it is a more a psychological concept than a physiological one. The first generation of questionnaires assessing body functionality pertain only to specific domains of body functionality. For example, the Physical Condition Subscale of the Body Esteem Scale (Franzoi & Shields, 1984) includes questions pertaining mainly to the physicality and sexualization of the body. The Self Objectification Questionnaire (Noll & Fredrickson, 1998) focuses on the physical objectification of women relative to physicality. The Body Surveillance Subscale of the Objectified Body Consciousness Scale (McKinley & Hyde, 1996) again targets body shame and appearance control in only women, ignoring other groups affected by the constructs. These scales, although making important contributions to the research of body functionality, do not capture body functionality in a holistic manner that goes beyond the physical appearance and abilities. This risks limiting body functionality assessment to able-bodied individuals (Alleva & Tylka, 2021).

The Functionality Appreciation Scale (FAS) has become the most widely used scale for the measure of body functionality (Alleva et al., 2017). Consisting of seven items, the FAS was designed to measure body functionality appreciation holistically, i.e., not specific to any one domain of body functionality (Alleva et al., 2017). Thus, items assess not only physical capability, but also internal processes, bodily perceptions, creative endeavors, and communication with others (Alleva & Tylka, 2021). Participants score the seven items on a range from 0 (strongly disagree) to 4 (strongly agree). An example of such an item is “*I appreciate my body for what it is capable of doing*.” Items are also designed to be all-inclusive regarding physical capacity, thereby including adults with physical disabilities (Alleva et al., 2017).

Since scales that measure body awareness and body image are used as outcome measures to evaluate the effectiveness of an intervention, it is imperative to make sure that researchers and clinicians have a baseline understanding of what ‘normal’ scoring looks like in healthy adults, and a valid way of looking at the scoring of the scales (Tennant & Conaghan, 2007). This is only possible if those scales are rigorously assessed. Determining the structural validity of a psychometric test ensures that the intended topic of measurement is the actual topic measured upon administration. Within this context, unidimensionality pertains to whether a scale is assessing a single construct or trait. Early studies investigating FAS have confirmed sex invariance through confirmatory factor analysis. They also demonstrated good internal consistency, test-retest reliability, and correlations between body image and well-being or the lack thereof (Alleva & Tylka, 2021). Exploratory and confirmatory factor analysis has been conducted in the US, Europe, Asia, and Australia (Alleva et al., 2017; Linardon et al., 2020; Namatame et al., 2022), and validated translations of the FAS are available in Malay, Romanian and Farsi, Japanese and Italian (Cerea et al., 2021; Namatame et al., 2022; Sahlan et al., 2022; Swami et al., 2019, 2021). However, the Cronbach’s alpha and exploratory or confirmatory factor analysis used in these studies assess scales based on their ordinal level of measurement. These measures assume that rating scales are linear and all items are agreeable, failing to take into account the potential variability of distances between the points, and item difficulty (Tennant & Conaghan, 2007).

The Rasch model is a probability model that states that if a person has a higher ability on a certain trait (e.g., functional appreciation; motor function), that person should have a greater probability of obtaining a higher score. Performing the Rasch analysis examines the unidimensionality of the scale and produces a map showing the hierarchy of items from easy to difficult, i.e., it is easier to obtain a higher score on the easy items than on the hard items. The underlying statistics used in the Rasch analysis are Chi squares, in which the observed scores are compared to the expected scores, matching the probability model. This analysis also converts the original ordinal scale to an interval scale. It places the ability of participants and the difficulty of items on one continuous scale, using the unit of a ‘logit’ (Wu, 2007). The scale will have good targeting if the average ability of the person on that trait matches the average difficulty of the item within a range of 0.5 logits (J. M. Linacre, n.d.-a). No previous study has conducted a Rasch analysis on the FAS. Because Rasch analysis converts an ordinal scale into an interval scale, the Rasch-based FAS will allow for future monitoring of precise, post-intervention changes in functional appreciation, given that the FAS is unidimensional and has good targeting and fit.

The development of a Rasch-based FAS creates an opportunity to investigate whether causality exists between variables, such as investigating the impact of body awareness training in adults with pain or mental health conditions and evaluating whether their functionality appreciation improves over time. Clinical body awareness training therapies involve establishing a connection between one’s mental and physical self (de Jong et al., 2016). Prior research provides evidence that Basic Body Awareness Therapy or BBAT, produces significant effects in areas such as chronic pain (Bergström et al., 2014), eating disorders, depression (Danielsson & Rosberg, 2015), and schizophrenia (Hedlund & Gyllensten, 2010), emphasizing the mental and physical benefits of the mind-body connection. Improvements in both physical and mental health, as well as body image and appreciation, are also reported with recreational activities that incorporate body awareness, including yoga (Halliwell et al., 2018; Impett et al., 2006; Rivest-Gadbois & Boudrias, 2019), Tai Chi and Qigong (James et al., 2021), dance (Burgess et al., 2006) and Pilates (Cruz-Ferreira et al., 2011). However, an assessment of the effect of yoga on functionality appreciation using the standard FAS failed to show any significant improvement in this measure (Alleva et al., 2020).

The aims of the present study are to (1) determine the structural validity of the FAS in community-dwelling adults in the US using the Rasch probability model, and (2) to investigate whether there is a significant difference in scoring between adults who currently perform body awareness training and those who do not.

## Materials and Methods

### Participants

Participants were recruited at the Minnesota State Fair and Highland Fest. All participants were 18-99 years of age and spoke English. Participants who were non-English speakers or pregnant were excluded from the study. General demographic information, as well as self-reports of pain or mental health conditions, were collected. They were asked about whether they ever did or were currently performing breathing exercises, mindfulness or relaxation exercises, or other body awareness training such as Qigong, Tai Chi, yoga, and martial arts. This demographic, clinical, and behavioral information allows us to determine if the scale is functioning differently for different groups based on those characteristics. Additional responses indicating participation in body awareness training also allowed us to examine the impact of body awareness training on FAS logit scores.

The Institutional Review Board of the University of Minnesota approved the study (IRB# STUDY00005849), and the study was performed in accordance with the Declaration of Helsinki. Since this was an anonymous survey where no identifying information was collected, consent was not signed but was acknowledged through the forms provided. Additionally, participants completed the University of California, San Diego Brief Assessment of Capacity to Consent (UBACC Assessment) as proof of their understanding of the consent (Jeste et al., 2007).

Participants unable to obtain a perfect score of 20 on the UBACC Assessment were excluded from the study. Healthy participants who had expressed interest in participating in research from the Brain, Body, Mind Lab, were invited to participate in this research project through an e-mail with a link to the questionnaire. Since no identifying information was collected, it was not possible to trace who responded to this request. In 2021, as part of a clinical trial, FAS results were also collected from a group of adults with spinal cord injury and neuropathic pain, approved by the IRB of the University of Minnesota (IRB# STUDY00008476), and their baseline scores were used for this analysis. They signed informed consent on the secure REDCap platform of the University of Minnesota.

### Outcome measure

The FAS has 7 items, ranging from 0 (strongly disagree) to 4 (strongly agree) with higher scores reflecting a higher level of appreciation for the functionality of the body.

### Statistical analysis

#### Rasch Analysis

The Rasch Measurement Theory evaluates structural validity: whether the scale is measuring one dimension (unidimensionality) and whether the items and the scale as a whole are fitting the probabilistic Rasch model. We used the statistical program Rasch Unidimensional Measurement 2030 Software (RUMM2030) to perform the Rasch analysis. We followed the Rasch Reporting Guideline for Rehabilitation Research (RULER) to report our results (Mallinson et al., 2022) (Van de Winckel et al., 2022).

The Rasch analysis produces several outcomes:

1. It verifies whether scoring categories for each item are fitting the probabilistic model, stating that a person with a higher ability in functional appreciation should score higher on an item than a person with a lower ability in appreciation of the body functionality. If observed scores on an item do not react in this way, then the Rasch analysis can show **reversed thresholds** for certain scoring categories, usually requiring a merge of scoring categories (i.e., rescoring of the items) (Pallant & Tennant, 2007).
2. **Overall fit, item fit, and person fit** are analyzed with Chi-square statistics to verify whether the observed scores match the expected scores of the probability model. Residuals greater than 2.5 indicate item misfit (Tennant & Conaghan, 2007).
3. The **person separation reliability** outcome measures how well we can differentiate high abilities from low abilities in a specific trait in persons (M. Linacre, n.d.). PSR ranges from 0-1.0, where a higher PSR indicates a better separation and more precise measurement (Wright & Stone, 1999). A score above 0.70 allows us to distinguish different abilities in groups; a score above 0.9 allows us to distinguish abilities in individual persons (Mallinson et al., 2014). The **mean error variance** is a type of standard error of measurement (J. M. Linacre, n.d.-b).
4. Good **targeting** is obtained when the average person location (in logits) is within a range of - 0.5 to +0.5 logits of the average item location, which by default is set at 0 logits (J. M. Linacre, n.d.-a). Moreover, floor and ceiling effects need to be reported when 15% or more participants have a minimum or maximum score on the scale (McHorney & Tarlov, 1995).
5. **Differential item functioning (DIF)** evaluates whether the hierarchy of items (in terms of difficulty of the item) is maintained across demographic, clinical, or behavioral variables. For example, men and women respond in the same way on this scale (Uddin & Islam, 2019). DIF is reported when the responses from one group are shifted more than 0.5 logits from the other group. The DIF investigated in this study were age (less than 65 years of age, vs 65 or older); sex (male, female, other); group (healthy adults, adults with self-reported mental health conditions, adults with pain); currently doing breathing exercises (yes, no); currently doing body awareness training (yes, no).
6. **Principal Component Analysis of Residuals (PCAR)** is an additional technique to investigate unidimensionality by extracting the common factor that explains the most residual variance under the hypothesis that there is such a factor. Ideally, the percentage total variance accounted for by the first principal component should be less than 10% with an eigenvalue of less than 2. The latter reflects that the variance is explained by 1 underlying trait. If this is not the case, then paired *t*-tests can be used between 2 subtests of items that load positively and negatively (with correlations smaller than -0.3 or larger than 0.3) on the first principal component, to investigate unidimensionality further. We have an additional indication that the scale is unidimensional if those paired *t*-tests report less than 5% significant differences in person locations on the two subtests (Tennant & Conaghan, 2007).
7. Residual correlations are reflecting a degree of **local item dependence (LID)**. This test examines whether two items have more in common with each other than with the whole scale. LID is reported when two items have a correlation at least 0.2 above the average residual item correlation (Christensen et al., 2017). In that case, it means that two items have more in common with each other than with the whole scale.

Bonferroni correction was applied for the statistical analyses that involved multiple comparisons.

### Body awareness training statistical analysis

As a secondary analysis, we first use descriptive statistics to report on the frequency of participants currently performing body awareness training. To examine the significant difference in FAS Rasch-based scoring between adults who currently perform body awareness training and those who do not, we use the Shapiro-Wilk test to assess the normal distribution of the data and, depending on the outcome, we used a 2-sample *t*-test or Mann-Whitney *U* test to compare the data between the two groups of adults practicing vs. not practicing body awareness-related modalities.

## Results

### Demographical, Clinical, and Behavioral Data

The final dataset used for the Rasch analysis contained 567 participants, including 286 healthy adults (age = 52.15 ± 17.5), 145 adults with self-reported mental health conditions (age = 46.88 ± 16.78), and 136 adults with self-reported pain (age= 54.82 ± 15.36), among which 14 were adults with SCI and SCI-related neuropathic pain, which occurred 1 to 45 years prior, at locations between the C4 and L1 vertebrae. Adults who had both self-reported pain and self-reported mental health conditions were counted in the group of adults with self-reported pain. **Table 1** presents the demographic, clinical, and behavioral data, and mean FAS scores (ordinal data) for all participants in their respective groups: 3.46±0.51 for healthy adults, 3.39±0.58 for adults with mental health conditions, and 3.19±0.58 for adults with pain. **Figure 1** illustrates the race and ethnicity distribution within the participants.

**Table 1.** Demographic, Clinical, and Behavioral Characteristics and mean FAS scores of Participants by Group

|  | Healthy<br>Adults<br>(n=286) | Adults with Mental<br>Health Conditions<br>(n=145) | Adults with<br>Pain<br>(n=136) |
| --- | --- | --- | --- |
| Age (years, mean±SD) | 52.15±17.59 | 46.88±16.78 | 54.82±15.36 |
| $\geq 65$ | 198 | 118 | 83 |
| $< 65$ | 117 | 34 | 53 |
| Sex |  |  |  |
| Female | 168 | 111 | 83 |
| Male | 117 | 34 | 53 |
| Other | 1 | 0 | 0 |
| Self-reported mental health conditions | 0 | 145 | 48 |
| Self-reported pain | 0 | 0 | 136 |
| Spinal cord injury | 0 | 0 | 14 |
| Ever done breathing exercises | 150 | 114 | 87 |
| Currently doing breathing exercises | 103 | 74 | 54 |
| Ever done mindfulness or relaxation exercises | 121 | 98 | 60* |
| Current mindfulness or relaxation exercises | 80 | 65 | 36* |
| Ever done body awareness training | 163 | 109 | 89 |
| Currently doing body awareness training | 88 | 55 | 39 |
| FAS score (mean ±SD, ordinal data) | 3.46±0.51 | 3.39±0.58 | 3.19±0.58 |

**Legend:** \*: Data not assessed in adults with spinal cord injury. Body awareness training included but was not limited to dance training, martial arts, Tai Chi, Qigong, yoga, Pilates, barre, and P.Volve exercises.

**Figure 1.**
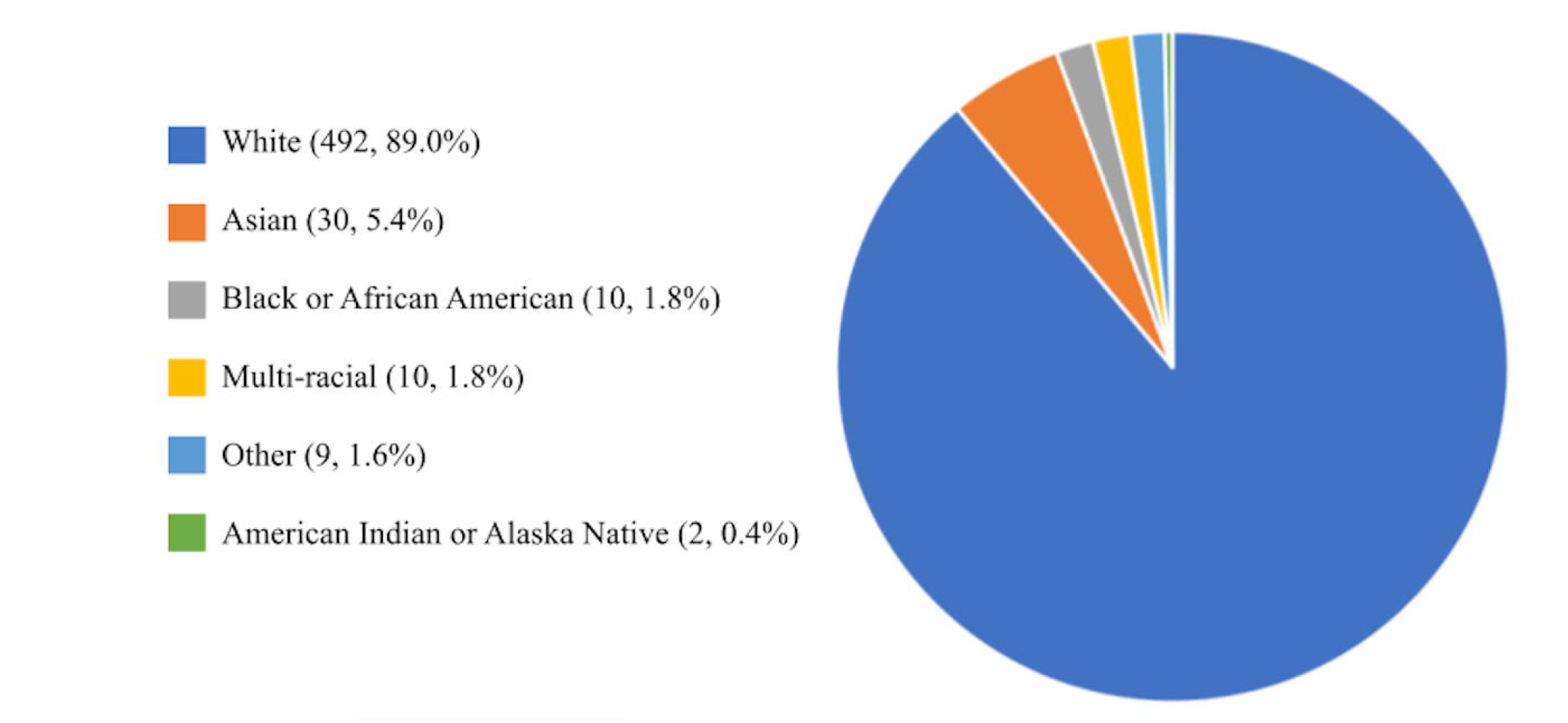
Race and Ethnicity Distribution. Pie chart showing race and ethnicity distribution of all 567 participants, demonstrating the lack of diversity in the group of community-dwelling adults.

### Rasch Measurement Analysis

The iteration analysis (**Table 2**) displays the Rasch analysis results of the person mean location, mean error variance, floor and ceiling effect, overall fit, item and person fit, number of items with disordered thresholds, number of misfitting items and persons, Principal Components Analysis of Residuals (PCAR), and Person Separation Reliability (PSR). Details of the iteration steps are presented below.

**Table 2.**
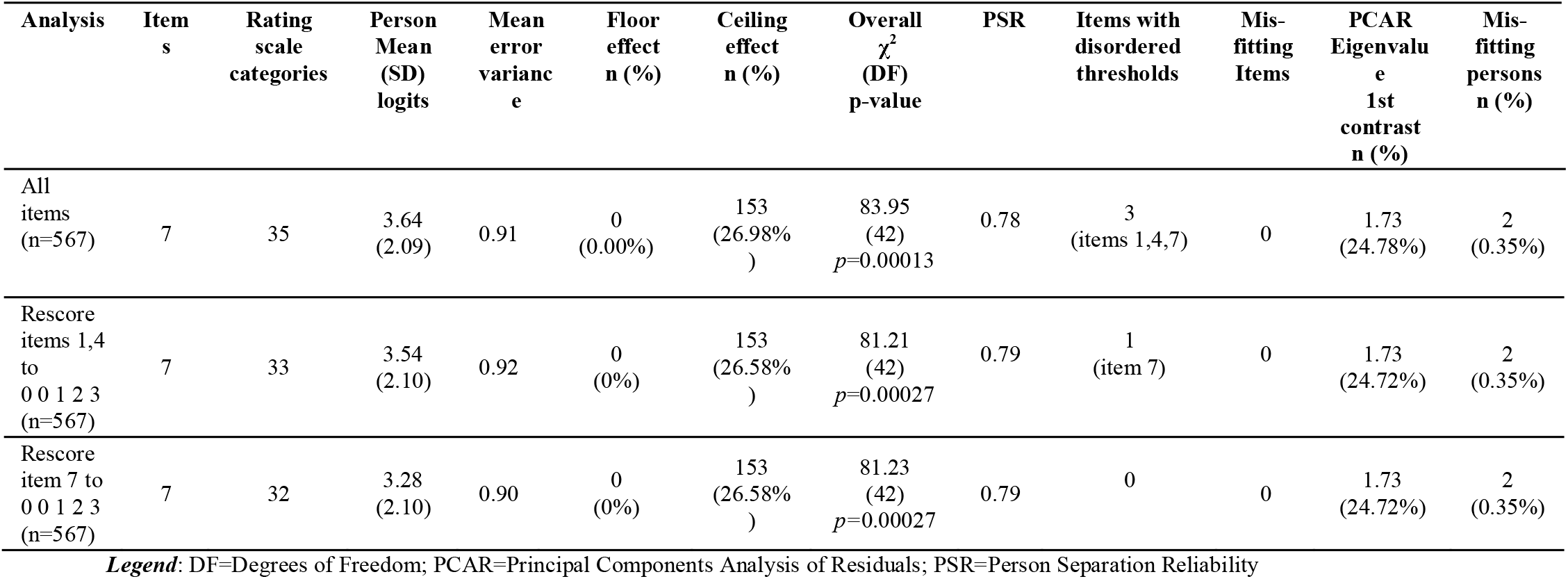
Iteration Analysis Table of the FAS

| Analysis | Items | Rating scale categories | Person Mean (SD) logits | Mean error variance | Floor effect n (%) | Ceiling effect n (%) | Overall $\chi^2$ (DF) p-value | PSR | Items with disordered thresholds | Mis-fitting Items | PCAR Eigenvalue 1st contrast n (%) | Mis-fitting persons n (%) |
| --- | --- | --- | --- | --- | --- | --- | --- | --- | --- | --- | --- | --- |
| All items (n=567) | 7 | 35 | 3.64 (2.09) | 0.91 | 0 (0.00%) | 153 (26.98%) | 83.95 (42) $p=0.00013$ | 0.78 | 3 (items 1,4,7) | 0 | 1.73 (24.78%) | 2 (0.35%) |
| Rescore items 1,4 to 0 0 1 2 3 (n=567) | 7 | 33 | 3.54 (2.10) | 0.92 | 0 (0%) | 153 (26.58%) | 81.21 (42) $p=0.00027$ | 0.79 | 1 (item 7) | 0 | 1.73 (24.72%) | 2 (0.35%) |
| Rescore item 7 to 0 0 1 2 3 (n=567) | 7 | 32 | 3.28 (2.10) | 0.90 | 0 (0%) | 153 (26.58%) | 81.23 (42) $p=0.00027$ | 0.79 | 0 | 0 | 1.73 (24.72%) | 2 (0.35%) |

**Legend:** DF=Degrees of Freedom; PCAR=Principal Components Analysis of Residuals; PSR=Person Separation Reliability

Three items were rescored because they contained reversed thresholds. Items 1 (“*I appreciate my body for what it is capable of doing*”) and 4 (“*I acknowledge and appreciate when my body feels good and/or relaxed*”) were rescored from [0 1 2 3 4] to [0 0 1 2 3]. Item 7 (“*I respect my body for the functions that it performs*”) was rescored in the next iteration from [0 1 2 3 4] to [0 0 1 2 3].

**Figure 2** shows the Rasch-based scoring sheet after rescoring. The items were renumbered to reflect the hierarchical order from the easiest item, “*I appreciate that my body allows me to communicate with others*”, at the top, to the most difficult item, “*I acknowledge and appreciate when my body feels good and/or relaxed*”, at the bottom. The scale demonstrated excellent person fit, as only 0.35% (2 participants out of 567) had a fit residual greater than 2.5.

**Figure 2.**
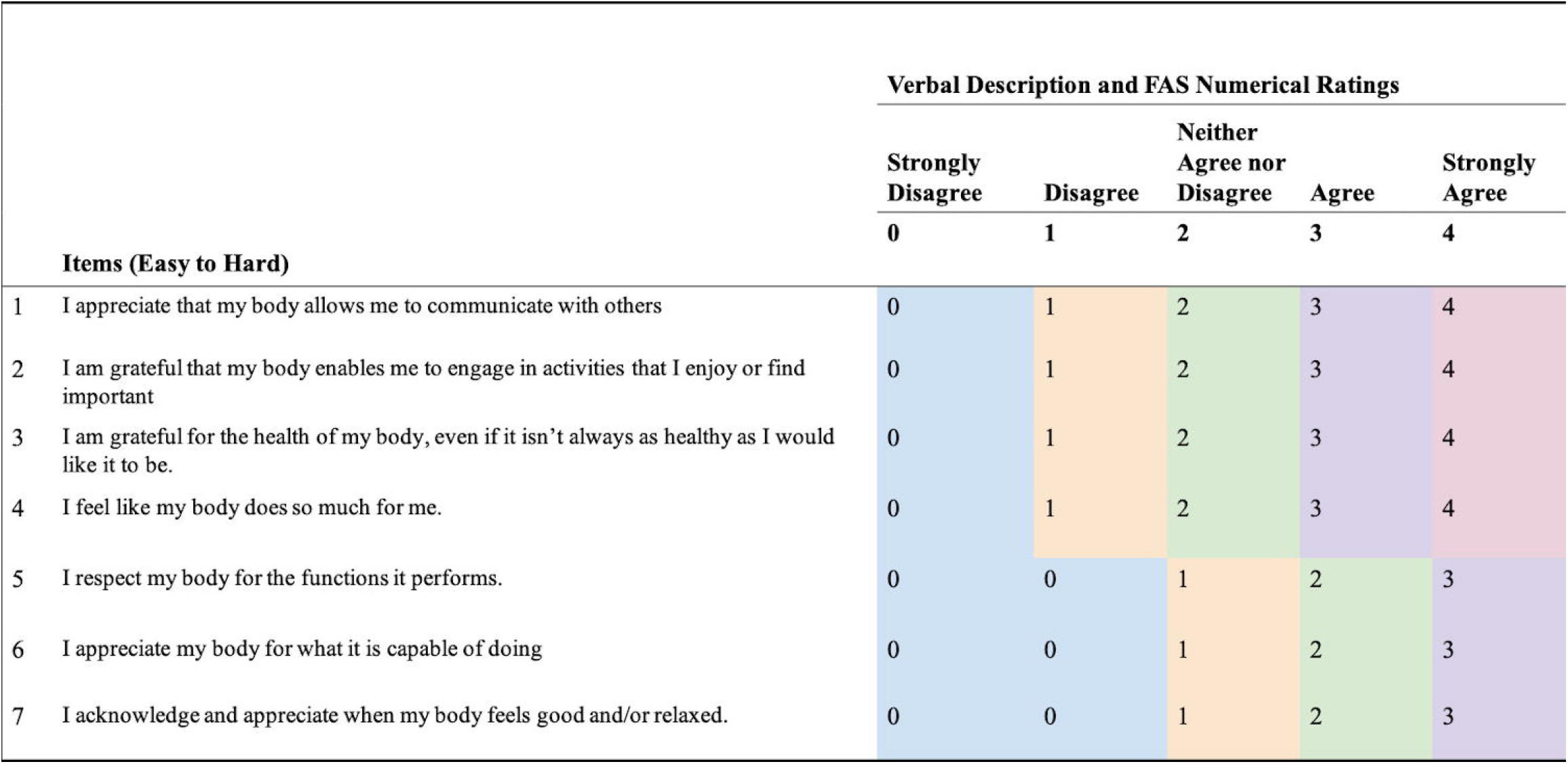
Rasch-Based FAS Scoring Sheet. Rasch-Based FAS Scoring Sheet is a representation of all the items with their respective scoring categories (ranging from strongly agree to strongly disagree). The colors indicate the rescoring of scoring categories of ‘strongly disagree’ and ‘disagree’ for items 5, 6 and 7.

There were no misfitting items (Table 3).

**Table 3.** Item Fit Statistics of the Rasch-based FAS Scale

| Item number | Item Difficulty | SE | $\chi^2$ | Item Fit | p-value |
| --- | --- | --- | --- | --- | --- |
|  | (Logits) |  |  | Residual |  |
| 1 | 0.82 | 0.10 | 13.73 | 0.02 | 0.03 |
| 2 | -0.18 | 0.09 | 4.51 | -1.26 | 0.61 |
| 3 | -1.33 | 0.10 | 3.42 | -1.27 | 0.76 |
| 4 | 0.83 | 0.09 | 15.63 | 1.90 | 0.02 |
| 5 | -0.42 | 0.09 | 11.54 | -3.33 | 0.07 |
| 6 | -0.05 | 0.08 | 10.04 | -2.13 | 0.12 |
| 7 | 0.32 | 0.09 | 22.37 | -5.83 | 0.001 |

**Legend:** SE: Standard Error; *p*-values are Bonferroni-corrected for multiple comparisons ( $\alpha$ corrected value at 0.007)

The item threshold map arranges items in order of difficulty (easiest to hardest; top to bottom) along a logit scale ranging from -7 to 5 with the scoring category thresholds for each item displayed (**Figure 3**). A logit scale not only represents the difficulty of the items but also the participants’ ability; a higher logit score indicates a high appreciation for the functionality of their body. The PSR value for the FAS was 0.79, which means that it is possible to differentiate groups of people according to their ability level of functional appreciation. There was no floor effect (0.00%), but there was a significant ceiling effect at 26.98% (143 participants out of 567). The person mean location of the whole group was 3.28±2.10 logits, meaning that the items are too easy for community-dwelling adults in the US. Even when the person distribution is categorized per group (i.e., healthy adults, adults with self-reported pain, adults with mental health conditions), the person mean location is still larger than 0.5 logits, ranging from 2.58±2.04 for adults with pain, to 3.35±2.19 for adults with mental health conditions, to 3.58±2.00 for healthy adults **(Figure 4**).

**Figure 3.**
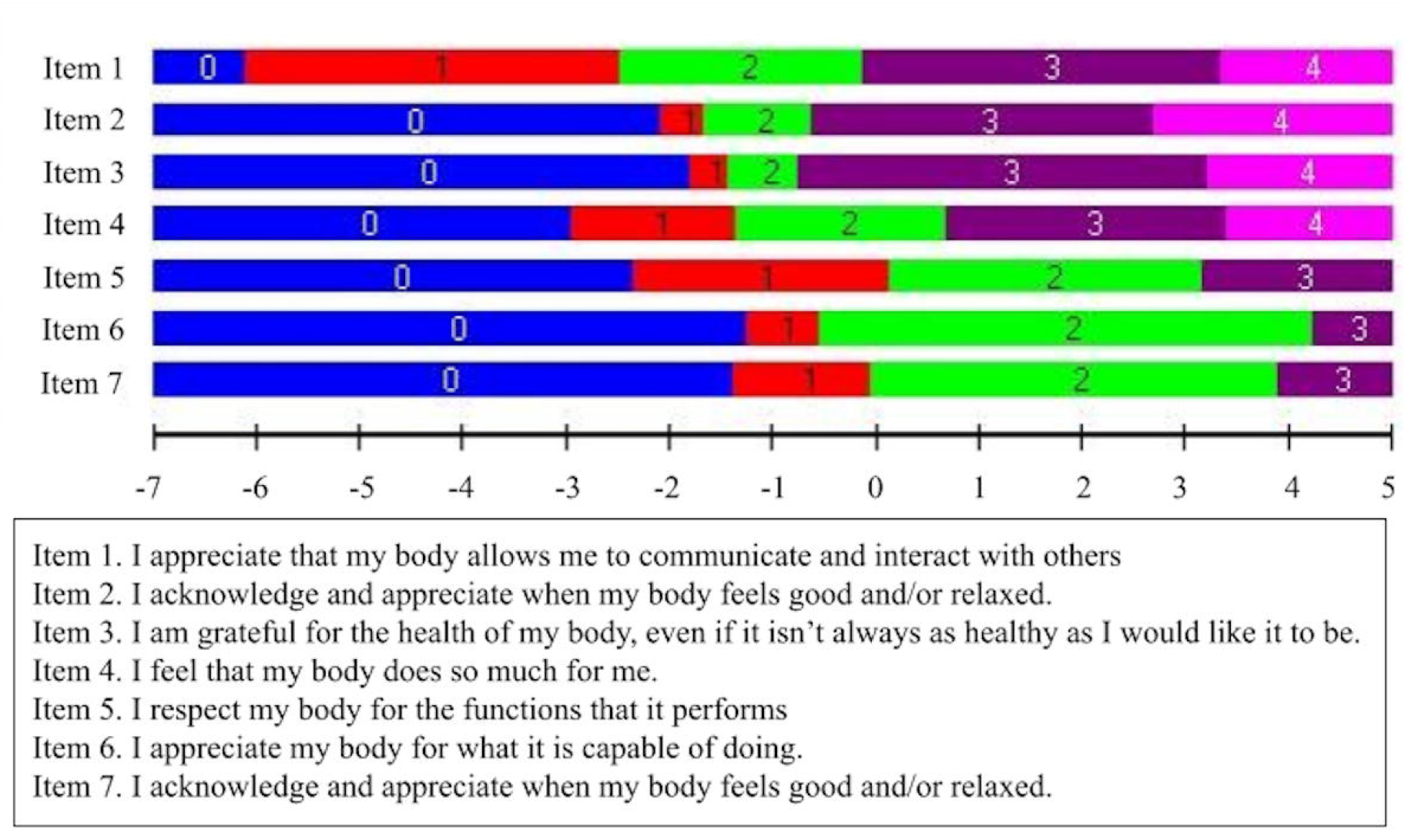
Item Threshold Map. The item threshold map shows items arranged in order of difficulty: easiest (“*I appreciate that my body allows me to communicate and interact with others*”) to hardest (“*I acknowledge and appreciate when my body feels good and/or relaxed”*). The horizontal ruler indicates the Rasch-based functionality appreciation score in logits corresponding to the scoring categories for each item.

**Figure 4.**
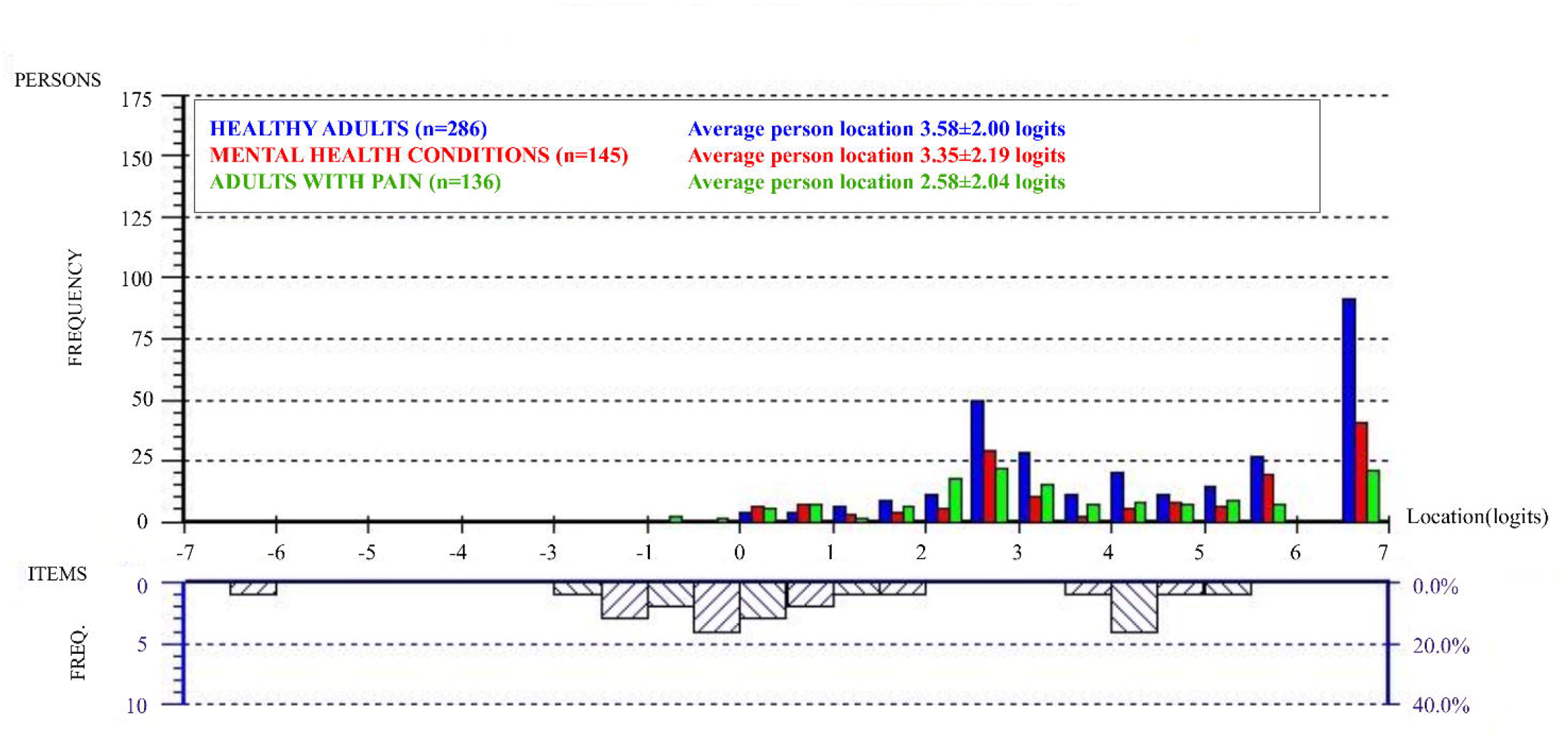
Person-Item Threshold Distribution. The person-item threshold distribution contains histograms that indicate the frequency of participants at different functionality appreciation ability levels (logit scores). The histograms are split and organized by demographic groups of healthy adults (blue), adults with mental health conditions (red) and adults with pain (green). The histogram below (chart with diagonal lines) indicates the frequency of item thresholds organized along the same logit scale (from easiest on the left to hardest on the right side of the ruler).

The PCAR eigenvalue was 1.73 and there was a percent variance of 24.72%. The paired *t*-tests revealed that only 3.35% of persons had significantly different person locations among the two subsets of items, thereby supporting the unidimensionality of this scale, namely (Tennant & Conaghan, 2007), measuring functionality appreciation.

There was no DIF found for any of our demographic, clinical, or behavioral variables, which had subgroups with n=200 or greater. Consequentially, item difficulty is working in the same way regardless of sex, subgroup allocation, or whether participants performed breathing or body awareness training. No consequential LID was found.

### Types of body awareness training

In this study, 32.1% of the participants reported currently performing some type of body awareness training, while 11.1% of the participants indicated participating in multiple modalities. The most practiced modality was Yoga, followed by Pilates, and dance training. The frequencies of adults in the community practicing the different types of body awareness are displayed in **Figure 5**.

**Figure 5.**
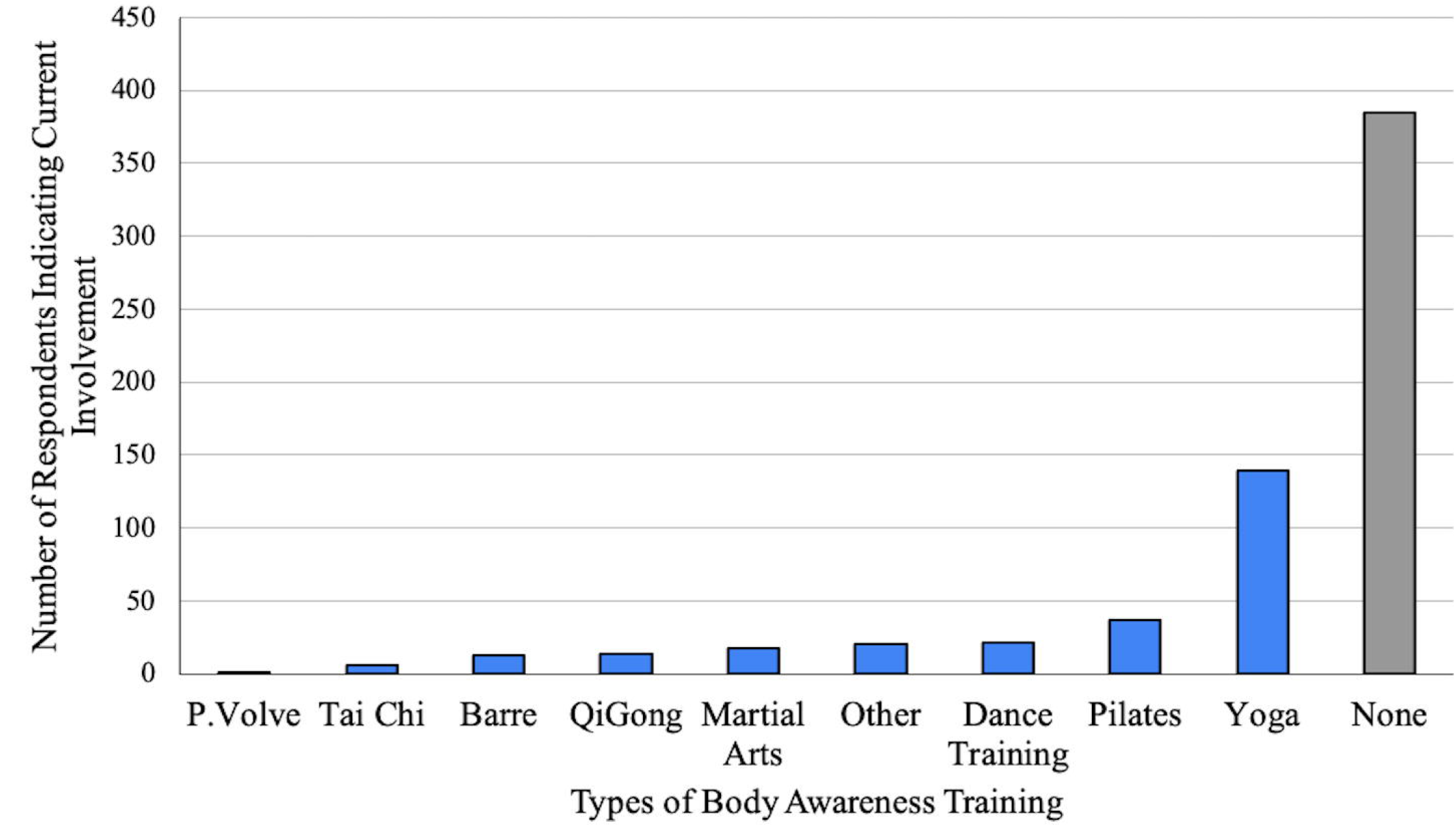
Body Awareness Training Distribution. The body awareness training frequency is presented as a bar chart specifying the frequency of participants who did not engage in certain types of body awareness training, or those who did perform such training. Those who did report this training are further separated out into specific types of body awareness training. P.Volve (founded in New York) is a low-intensity exercise program.

### The difference in FAS Rasch-based scoring between adults who currently do body awareness training vs. those who do not

The Shapiro-Wilk test demonstrated that the data was not normally distributed (*W* = 0.85, *p* < .0.0001). The group of participants who reported not currently performing body awareness exercises (n=385) had a significantly lower average location (in logits) than the group of participants who currently performs body awareness training (n=182, *U* = 4.33, *p* < 0.0001). The median and Interquartile Range (IQR) score of the group that reported not practicing body awareness was 2.92 logits (IQR = 3.15) and the group that reported practicing body awareness was 3.72 logits (IQR=4.06; **Figure 6**).

**Figure 6.**
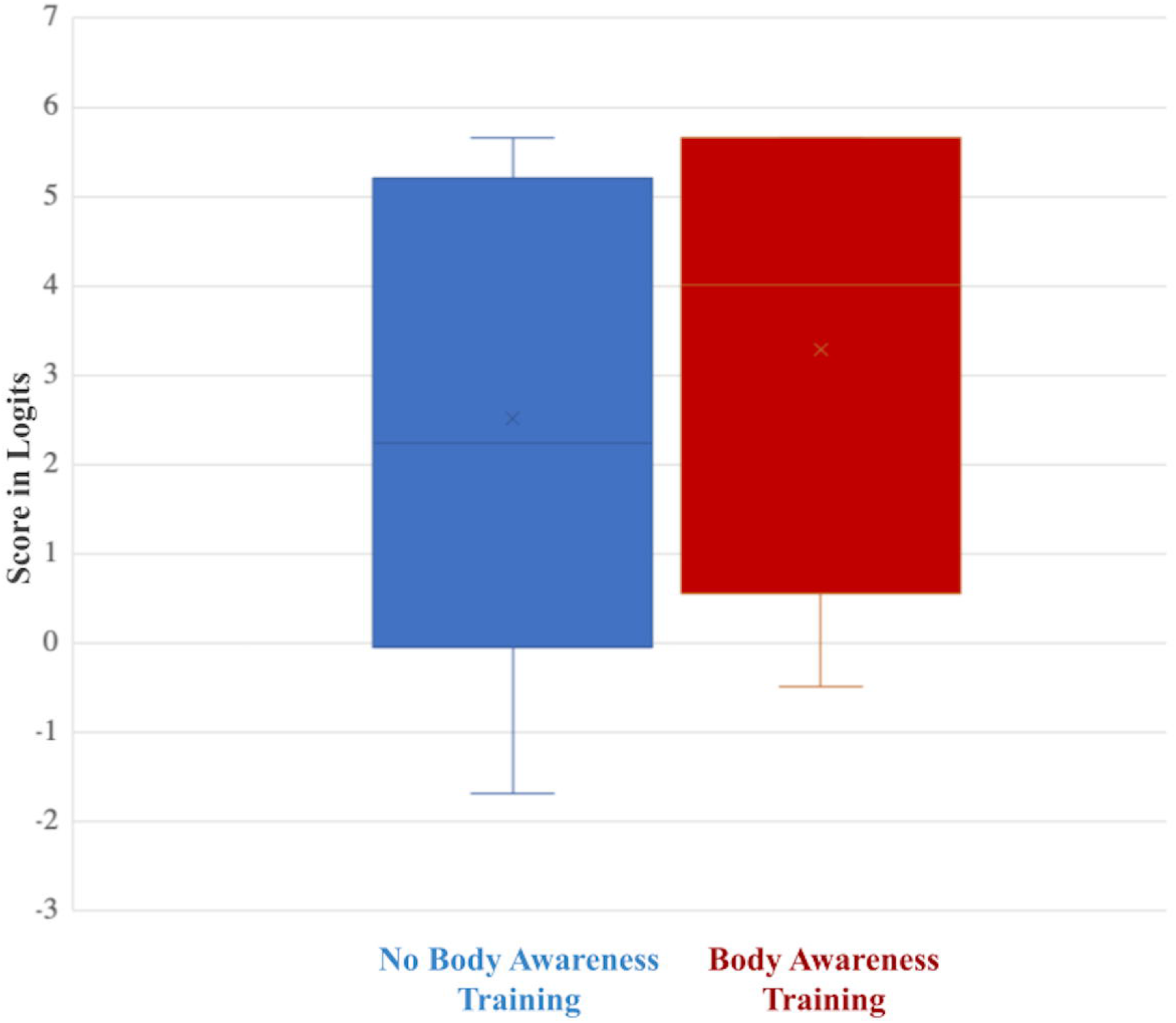
Score in Logits of Body Awareness Training participants vs. non-participants. Boxplots present mean logit locations, including median and interquartile ranges for individuals reporting no body awareness training (blue; n=385) and reporting body awareness training (red; n=182)

## Discussion

Rasch validation of the FAS demonstrated that the FAS is unidimensional and has a good item and person fit. The finding of unidimensionality agrees with previous validation studies on the FAS that employed exploratory-to-confirmatory factor analyses (CFA; Cerea et al., 2021; Namatame et al., 2022; Sahlan et al., 2022; Swami et al., 2021). The Rasch analysis also allowed for specific analysis on the item and person fit, which has not been previously measured. These results remained supportive of the CFA-based findings from other studies that confirmed the validity of this scale.

In accordance with the original study and subsequent validation studies, this research assessed the FAS in a non-clinical setting (Alleva et al., 2017). A significant ceiling effect and positive person mean location revealed that the items were too easy for community-dwelling adults. Although it has not been previously discussed, this result is consistent with the original paper reporting mean FAS ordinal scores in community-dwelling adults ranging from 3.88 to 4.24 out of a maximum score of 5 (Alleva et al., 2017). The lowest mean FAS scores reported previously, in the validation of a Japanese translation, was 3.60 out of 5 (Namatame et al., 2022). Within the present study, data also showed mean FAS scores of 3.38±0.56. These are among the lowest FAS scores reported, yet, when transformed to the logit scale with Rasch analysis, the FAS was still too easy for them as well.

Using the Rasch-based FAS, we found a significant increase in functionality appreciation in individuals who performed body awareness training as compared to those who did not. Another cross-sectional study found that athletes had higher mean functionality appreciation scores than non-athletes on the ordinal scale outcome (Soulliard et al., 2019). Combined with our results, this may indicate that consistent athletic training could also support a higher functionality appreciation.

Two limitations to this study are worth noting. First, Adults with SCI and neuropathic pain were combined with the other adults who self-reported pain because the sample size was too small to identify fit to the model for this group separately. Further studies are needed to identify the targeting of the FAS specifically for adults with SCI and neuropathic pain. Second, the state of Minnesota has an 82.85% White population and a 6.41% Black or African American population (*Minnesota Population 2022 (Demographics, Maps, Graphs)*, n.d.). Therefore, 89.0% of our respondents were White, and only 1.8% were Black or African American. As a result, our findings were not representative of all racial and ethnic groups. Further studies should investigate the targeting of FAS in more inclusive groups.

In conclusion, functionality appreciation represents an important new facet in improving overall body image that is not based on physical appearance (Alleva et al., 2017). While the Rasch analysis of the FAS demonstrates good item and person fit, the targeting needs to be addressed to improve the effectiveness of FAS in assessing community-dwelling adults. With a validated test, there are many directions to explore the concept of functionality appreciation. We found that those that performed body awareness training scored higher on functional appreciation. Future studies should also expand on how specific social media platforms impact body functionality appreciation scores for teenagers and young adults. Additionally, with the use of a valid functionality appreciation scale, we can explore how these perceptions could be linked to individual traits and identities. Future research should also test the scale’s validity among more diverse populations and in specific groups, such as adults with spinal cord injury and neuropathic pain. Once the psychometrics of the Functionality Appreciation Scale are good, it can be a powerful tool to help us assess our perception of viewing ourselves and the functionality of our bodies.

## Data Availability

Upon acceptance of the paper, the data will be made available on the Dryad data repository (data link below)

https://doi.org/10.5061/dryad.66t1g1k4d

## CRediT authorship contribution statement

**Sarah Feng\***: Data curation, Visualization, Writing - original draft; **Sydney McDaniel\***: Data curation, Visualization, Writing - original draft; **Ann Van de Winckel:** Conceptualization, Methodology, Formal analysis, Visualization, Writing - review & editing.

*These authors contributed equally to this work.

## Acknowledgements

The authors thank all participants and volunteers at the Minnesota State Fair. We would like to express our profound thanks to Dr. Kati Kragtorp and Marc Noël for the critical review of the manuscript.

## Funding

Dr. Van de Winckel’s research was supported by the National Institutes of Health’s National Center for Advancing Translational Sciences, grant UL1TR002494. The content is solely the responsibility of the authors and does not necessarily represent the official views of the National Institutes of Health’s National Center for Advancing Translational Sciences. The funders had no role in the study design, data collection, and analysis, decision to publish, or preparation of the manuscript.

## Declaration of Competing Interest

The authors report no competing interests.

## Notes

### Competing Interest Statement

The authors have declared no competing interest.

### Funding Statement

The research was supported by the National Institutes of Health National Center for Advancing Translational Sciences, grant UL1TR002494. The content is solely the responsibility of the authors and does not necessarily represent the official views of the National Institutes of Health National Center for Advancing Translational Sciences. The funders had no role in the study design, data collection, and analysis, decision to publish, or preparation of the manuscript.

### Author Declarations

The Institutional Review Board of the University of Minnesota approved the study (IRB# STUDY00005849), and the study was performed in accordance with the Declaration of Helsinki.

